# Identifying clusters of people with Multiple Long-Term Conditions using Large Language Models: a population-based study

**DOI:** 10.1101/2025.02.14.25322277

**Authors:** Alexander Smith, Thomas Beaney, Carinna Hockham, Bowen Su, Paul Elliott, Laura Downey, Spiros Denaxas, Payam Barnaghi, Abbas Dehghan, Ioanna Tzoulaki

## Abstract

**Background:** Identifying clusters of people with similar patterns of Multiple Long-Term Conditions (MLTC) could help healthcare services to tailor management for each group. Large Language Models (LLMs) can utilise complex longitudinal electronic health records (EHRs) which may enable deeper insights into patterns of disease. Here, we develop a pipeline, incorporating an LLM, to generate gender-specific clusters using clinical codes recorded in EHRs.

**Methods:** In this population-based study, we used EHRs from individuals aged ≥50 years from Clinical Practice Research Datalink in the UK. Longitudinal sequences of medical histories including diagnoses, diagnostic tests and medications were used to pre-train an LLM based on DeBERTa. The LLM, called EHR-DeBERTa, includes embedding layers for age of diagnosis, calendar year of diagnosis, gender, and visit number with a diagnosis vocabulary of 3776 tokens, covering the entire ICD-10 hierarchy. We fine-tuned EHR-DeBERTa using contrastive learning and generated patient embeddings for all individuals. A bootstrapping clustering pipeline was applied separately for females and males and gender-specific patient clusters were characterised by disease prevalence, ethnicity and deprivation.

**Findings:** A total of 5,846,480 patients were included. We identified fifteen clusters in females and seventeen clusters in males, grouped into five categories: i) low disease burden; ii) mental health; iii) cardiometabolic diseases; iv) respiratory diseases, and v) mixed diseases. Cardiometabolic and mental health conditions showed the strongest separation across clusters. People in low disease burden and mental health clusters were younger, whereas those in cardiometabolic clusters were older, with females in cardiometabolic clusters older than their male counterparts.

**Interpretation:** Using an LLM applied to longitudinal EHRs, we generated interpretable and gender-specific clusters of diseases, providing insights into patterns of diseases. Extending these methods in future to incorporate clinical outcomes could enable identification of high-risk patients and support precision-medicine approaches for managing MLTC.

## Introduction

Worldwide, a growing number of people are living with Multiple Long-Term Conditions (MLTC), a health state characterised by the co-existence of two or more chronic conditions.^1,2^ This presents a significant challenge for health systems,^3^ as those with MLTC experience worse health outcomes,^4^ poorer quality of life^5^ and greater use of health services.^6^ Furthermore, health services are often designed around the care of single diseases, resulting in an accumulating burden of clinical appointments for patients and challenges in navigating healthcare.^7^

One of the biggest difficulties in designing interventions to address the challenges of MLTC is clinical heterogeneity. Many different chronic conditions may be included in the definition of MLTC^8^ and there are many possible unique combinations of diseases that may occur in people.^9^ As a result, there is growing interest in identifying patterns of MLTC which commonly occur in a population.^10^ Clustering offers a data-driven approach to identifying these patterns, which generates groups of similar diseases, or of people based on similar patterns of diseases. Most clustering approaches have been applied to diseases, identifying strong patterns of cardiometabolic and of mental health conditions,^11,12^ but relatively few studies have identified individual clusters based on a holistic view of their comorbidities. Understanding groups of people with similar conditions may more directly inform understanding of shared causes and outcomes, and inform health service design, such as designing a service tailored to a specific cluster.^13,14^

Moreover, studies in MLTC have often employed cross-sectional designs, capturing the diseases a person has at a single point in time. However, this does not account for the temporal order of disease development, which has been found to have significant value in predicting healthcare utilisation and clinical outcomes.^15^ Use of deep learning models, such as transformers, enable sequences to be incorporated, and have been increasingly applied to predictive tasks in healthcare.^15–17^ These approaches can be applied to the rich information recorded within electronic health records (EHRs), which include symptoms, clinical examination findings, prescribed medications and laboratory test results, allowing the separation of phenotypes according to these factors and not only on diseases. To our knowledge, such approaches have not been applied to develop clustering methods to explain common patterns of diseases.

In this study, we aimed to generate gender-specific clusters of patients with similar sociodemographic profiles and sequences of clinical information recorded in EHRs, using a large and nationally representative sample of people registered to general practices (GPs) in the United Kingdom (UK). To achieve this, we first develop a transformer architecture which incorporates information on the chronological order of disease development, followed by clustering. Finally, we describe the resulting clusters by gender, socio-demographic profiles and diseases to identify common MLTC patterns within the population.

## Methods

### Data sources

This retrospective cohort study used data from Clinical Practice Research Datalink (CPRD) GOLD, a nationally representative dataset of approximately 10 million patients registered to GPs in the UK.^18^ CPRD contains comprehensive structured data from EHRs of registered individuals from contributing GP practices which includes diagnoses, prescriptions, laboratory test results and referrals. Data are also linked to a composite measure of socioeconomic deprivation, the Index of Multiple Deprivation (IMD) at patient level, which is grouped into quintiles.^19^ Secondary care records are also available for individuals eligible for linkage to Hospital Episode Statistics (HES) and Office of National Statistics (ONS) which includes diagnoses and laboratory test results for hospitalised patients, and death registration data. CPRD GOLD defines the gender (male or female) of an individual as recorded by the healthcare provider. The documented ethnicity of individuals was identified using SNOMED CT codes indicative of ethnicity within CPRD for individuals (Supplementary Table 1) with at least one ethnicity code. CPRD has established quality assurance through validation processes and the inclusion of GP practices meeting predefined research standards.^18^

### Eligibility

We included all individuals aged 50 years and over who were registered to a participating GP practice for at least three years on 31^st^ December 2022.

### Disease definitions

We included 54 chronic conditions and three risk factors within our definition of MLTC which were selected based on the combined list from the Global Burden of Disease Study and a recent Delphi consensus study on MLTC.^20,21^ We used available ICD-10 code lists from these studies (Supplementary Table 2) and refined each code list to better capture clinically relevant or ambiguous conditions. For CPRD data, which uses SNOMED-CT codes, ICD-10 codes were mapped to SNOMED CT using the 2022 release of SNOMED-to-ICD map by the NIH Unified Medical Language System to generate a SNOMED CT codelist for each chronic condition and risk factor.^22^ Therefore, clinical diagnoses were defined by the presence of either an ICD-10 or SNOMED CT for each condition.

### Longitudinal patient sequences of electronic health records

The study outline for generating a patient embedding, which is the high-dimensional vector that represents a patient’s medical history and MLTC patterns, is shown in Figure 1. For each patient, a longitudinal sequence of their medical history was constructed using clinical diagnoses, symptoms, medication prescriptions and laboratory test results. Further details on the sequence construction are given in the appendix.

**Figure 1.**
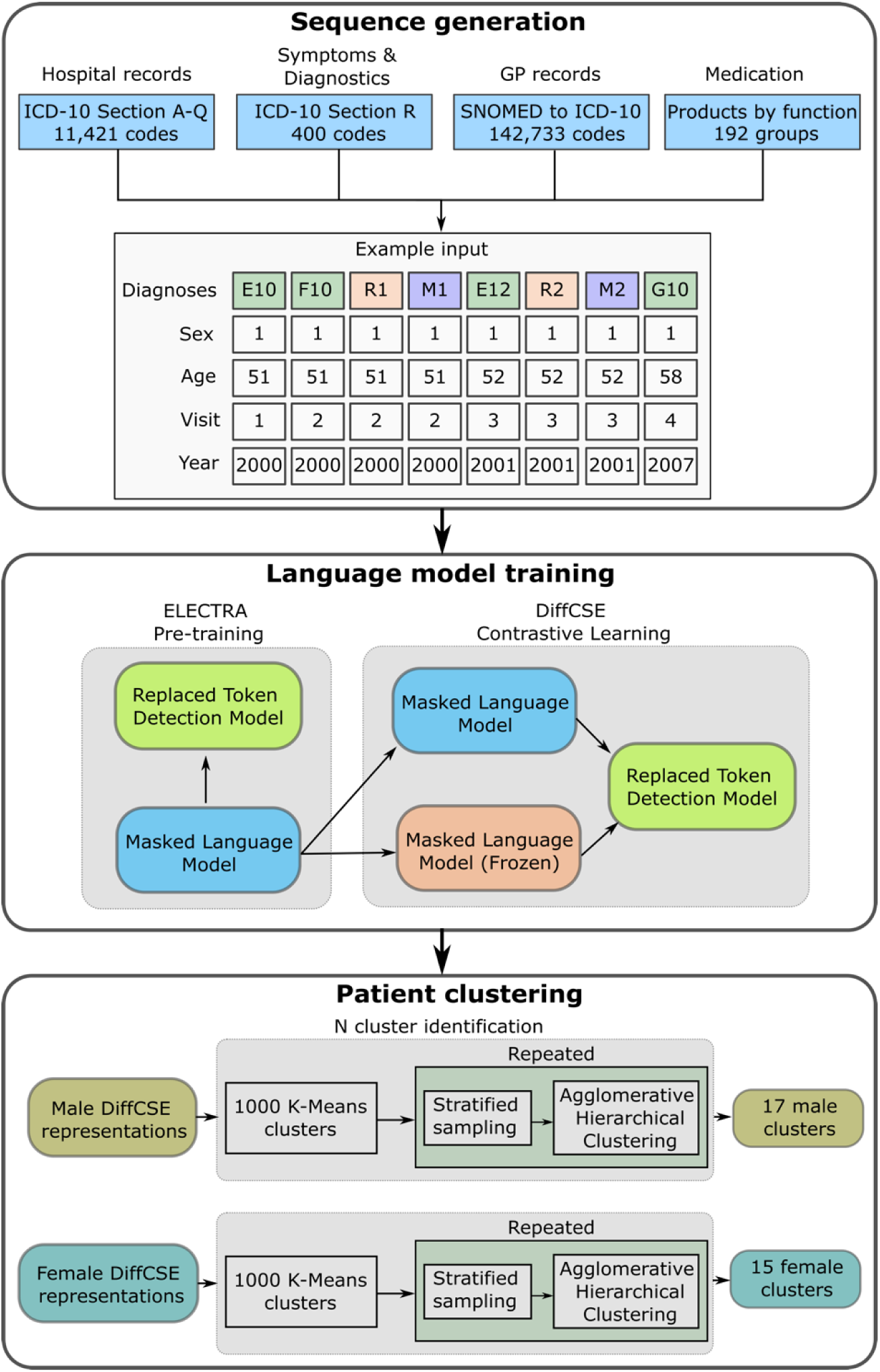
Study overview of training the large language model and generating patient clusters using EHRs

### Generating patient representations

To generate a quantitative representation of each patient’s sequence as an input for clustering, we developed a transformer architecture, named EHR-DeBERTa. Previous work applied to structured diagnostic codes in EHR data have used BERT (Bidirectional Encoder Representations from Transformers) architectures, which are a transformer-based deep learning models used for natural language processing.^15–17^ Here, we utilise a Decoding-enhanced BERT with disentangled attention (DeBERTa) architecture, which employs disentangled attention to represent words as two separate vectors (content and position) along with absolute word position embeddings, shown to perform better than the original BERT models in learning more complex relationships in several language modelling tasks.^23^ To improve the model’s ability to learn the relationships between events, we included embedding layers for patient age and calendar year at each event, gender of the individual, and visit number of the event.^15^ To train the EHR-DeBERTa model we utilised the same ELECTRA-style pre-training published with DeBERTa v3, followed by fine-tuning using Difference-based Contrastive learning for Sentence Embeddings (DiffCSE) to ensure robust and improved patient representations.^24,25^ Patient embeddings were generated for the entire cohort by feeding each longitudinal patient sequence, including all paired covariate sequences, through the DiffCSE fine-tuned EHR-DeBERTa model to generate a 786-dimensional vector representation for each patient. Further details of our pre-training and fine-tuning, along with our hyperparameters are given in the appendix.

To check that the model was learning clinically meaningful relationships between conditions, we evaluated the outputs with reference to clinically established disease relationships produced in earlier work.^26^ We did this by calculating the cosine similarity between pairs of clinically established co-morbidities (Supplementary Table 4) within the clinical code input embeddings of the fine-tuned model. Further details are given in the appendix.

### Generating gender-specific patient clusters

To generate gender-specific patient clusters, we stratified the cohort by gender and applied the clustering pipeline to each stratum separately. Robust identification of the optimal number of clusters for each gender was performed by bootstrapping agglomerative hierarchical clustering on 1% subsamples of each stratum across 500 repeats. To ensure subsampling was representative of the entire strata cohort, before subsampling we clustered the entire cohort into 1000 K-Means clusters and for each bootstrapping step we utilised these clusters with random stratified sampling to sample equally from each cluster. Optimal cluster numbers were identified using the mean and standard deviation silhouette score across all replicates.^27^ K-Means clustering using the optimal number of clusters was then applied to each stratum to identify gender-specific clusters.

### Cluster evaluation

We defined the clusters in terms of clinical and socio-demographic factors by testing the statistical association of cluster assignment. Associations with continuous variables were tested using 1-vs-all t-tests, binary variables were tested using Chi-squared tests. In addition, we calculated cluster specific disease-frequency – inverse patient frequency (c-DF-IPF), a method adapted from the natural language processing method of term-frequency – inverse document frequency (TF-IDF),^28^ to create cluster specific weighted disease frequency. DF-IPF reduces the weight of common diseases within the population and emphasises the diseases that are frequent within a cluster, but rare across all clusters. This allows better identification of the important diseases for a cluster based on local (cluster) importance and global (population) infrequency. More information is given in the appendix. Clusters were named and grouped clinically according to the common patterns of diseases within them, focusing on those conditions that varied significantly between clusters. Within each category, clusters were then numbered in descending order of prevalence.

We used Python version 3.11.5 and Pytorch version 2.0.1 to create and train the LLM models. Statistical analysis and data processing were performed using sci-kit learn version 1.3.2. Ethical approval for CPRD data and linkage to HES data was granted by CPRD’s Research Data Governance Process on 23rd September 2021 (protocol reference: 20_000209).

## Results

### Cohort description

A total of 5,846,480 patients were included in the analysis. The mean (standard deviation (SD)) age was 66 (16) years, 3,201,230 (54·8%) were female and 2,645,250 (45·2%) were male (Table 1). Information on ethnicity was recorded for 37·7% of participants, of whom 2,055,170 (93·4%) were White, 43,345 (2·0%) Black, 63,972 (2·9%) South Asian, 11,447 (0·5%) Mixed, and 26,097 (1·2%) of ‘Other’ ethnicity. Relatively more patients were living in less deprived areas, with 22·6% in the least deprived and 15·8% in the most deprived areas.

**Table 1:** Characteristics of the study population.

|  | Female | Male | Total |
| --- | --- | --- | --- |
| **N** | 3,201,230 (54.8%) | 2,645,250 (45.2%) | 5,846,480 |
| **Age (Median, IQR)** | 67 (54-81) | 66 (54-77) | 66 (54-79) |
| **Ethnicity (n, %)** | 1,189,387 (37.2%) | 1,010,644 (38.0%) | 2,200,031 (37.7%) |
| White | 1,110,639 (93.4%) | 944,531 (93.5%) | 2,055,170 (93.4%) |
| Black | 24,536 (2.1%) | 18,809 (1.9%) | 43,345 (2.0%) |
| South Asian | 33,212 (2.8%) | 30,760 (3.0%) | 63,972 (2.9%) |
| Mixed | 6,446 (0.5%) | 5,001 (0.5%) | 11,447 (0.5%) |
| Other | 14,554 (1.2%) | 11,543 (1.1%) | 26,097 (1.2%) |
| **Deprivation (n, %)** | 1,638,451 (51.2%) | 1,359,107 (51.4%) | 2,997,558 (51.3%) |
| 1 (Least deprived) | 372,275 (22.7%) | 304,134 (22.4%) | 676,409 (22.6%) |
| 2 | 366,841 (22.4%) | 299,736 (22.1%) | 666,577 (22.2%) |
| 3 | 347,845 (21.2%) | 285,278 (21.0%) | 633,123 (21.1%) |
| 4 | 299,559 (18.3%) | 248,905 (18.3%) | 548,464 (18.3%) |
| 5 (Most deprived) | 251,931 (15.4%) | 221,054 (16.3%) | 472,985 (15.8%) |

### Disease patterns within clusters

We identified an optimal number of 15 clusters in females and 17 clusters in males. Cardiovascular and metabolic diseases, mental health conditions and risk factors including hypertension, smoking, and obesity varied significantly in prevalence across clusters in both females (Figure 2) and males (Figure 3). Few diseases showed little variation across clusters. For example, there was no statistically significant difference in the prevalence of Addison disease across clusters in males or females. We found five broad groupings of the 15 female and 17 male clusters characterised by the following patterns: i) low disease burden (characterised lower than expected prevalence in all but 2 diseases); ii) mental health; iii) cardiometabolic diseases; iv) respiratory diseases and v) mixed diseases.

**Figure 2:**
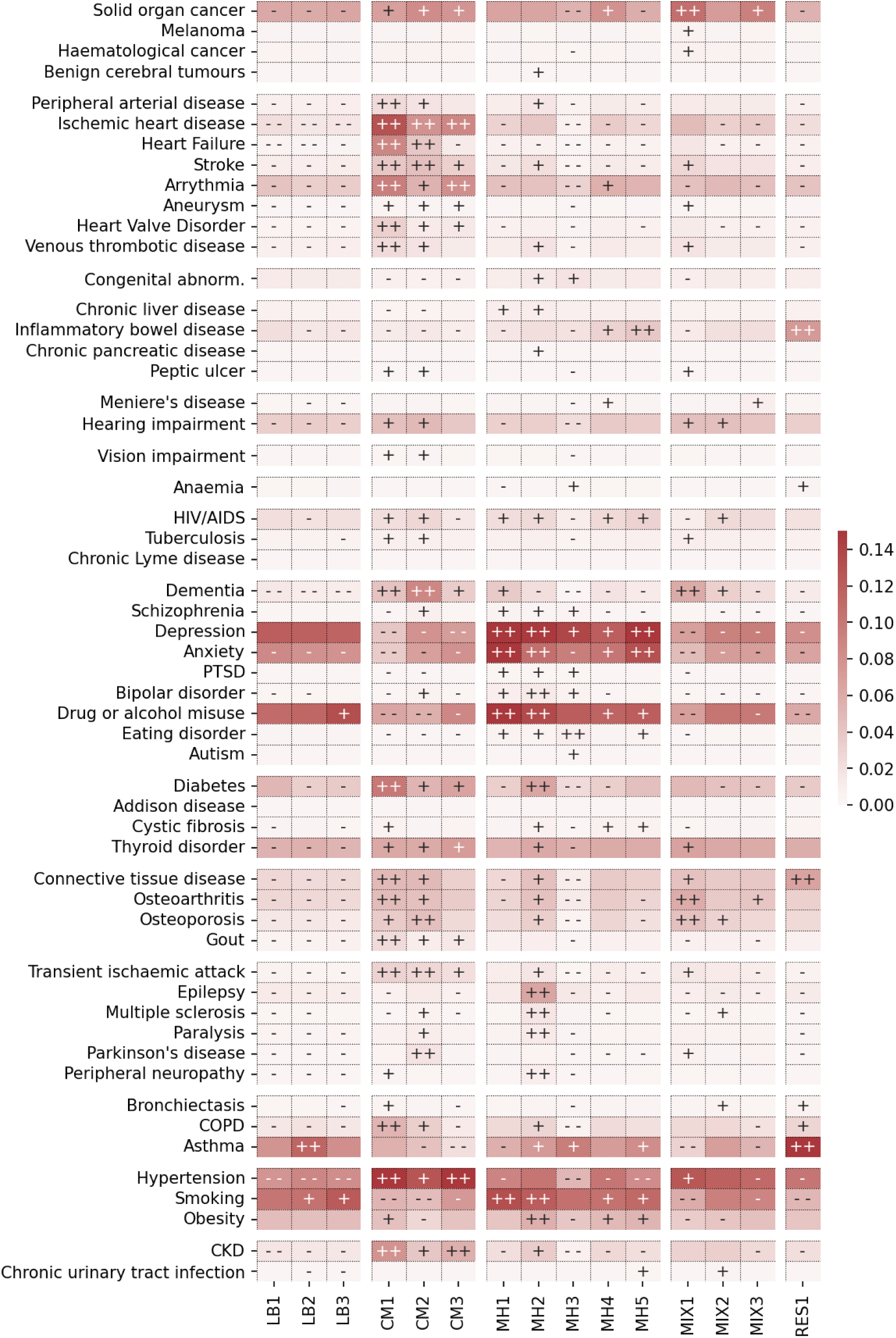
Disease prevalence across clusters of 3,201,230 females from CPRD, shown by cluster specific weighted frequency (c-DF-IPF) and symbols indicating the Z-score differences from Chi-squared tests (++ ≥ 50, 50 > + ≥ 10, - - ≤ −50, −50 < - ≤ −10). LB: low burden, CM: cardiometabolic, MH: mental health, MIX: mixed, RES: respiratory

**Figure 3:**
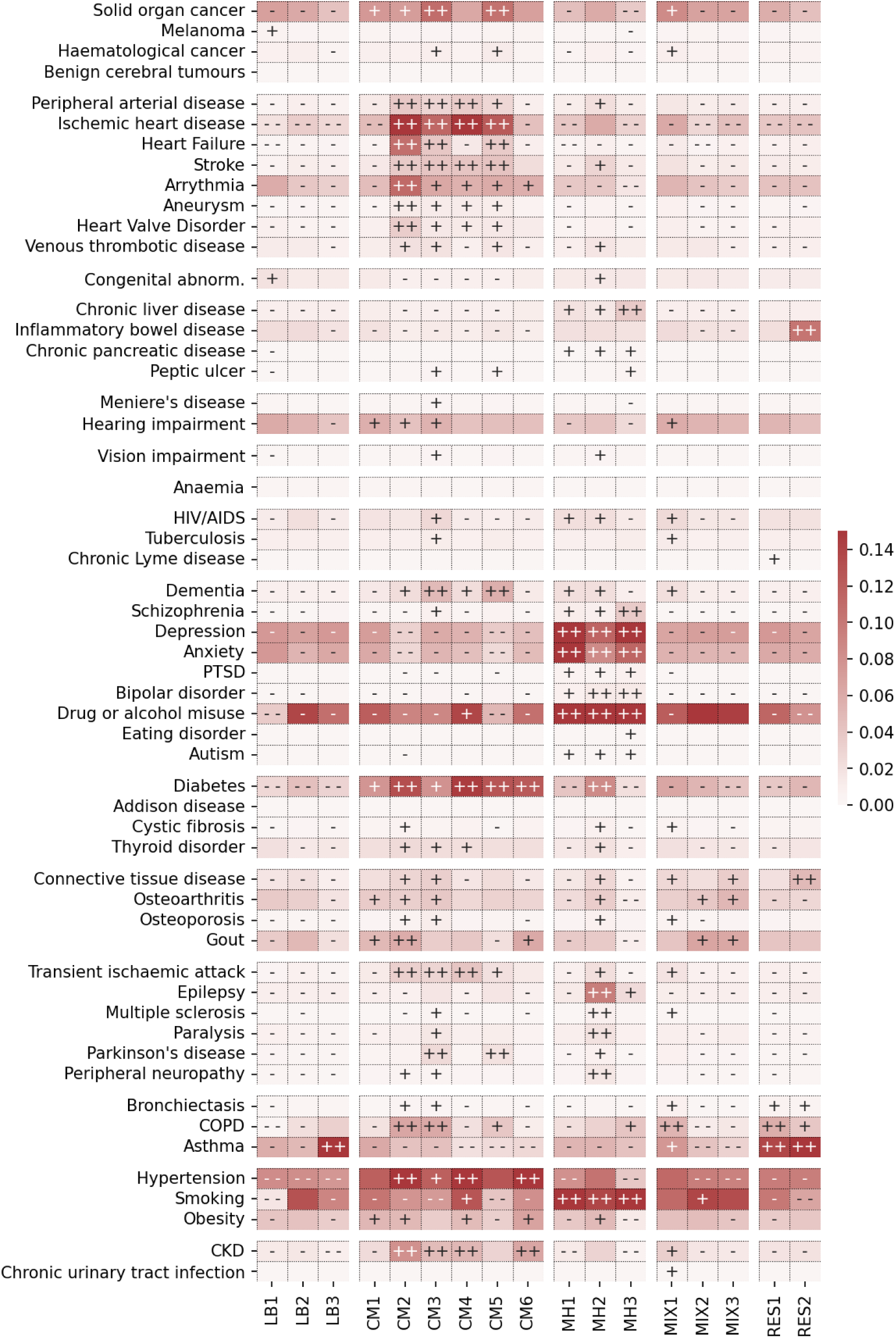
Disease prevalence across clusters in 2,645,250 males from CPRD, shown by cluster specific weighted frequency (c-DF-IPF) and symbols indicating the Z-score differences from Chi-squared tests (++ ≥ 50, 50 > + ≥ 10, - - ≤ −50, −50 < - ≤ −10). LB: low burden, CM: cardiometabolic, MH: mental health, MIX: mixed, RES: respiratory

In females (Figure 2), three clusters of relatively healthy individuals with low disease burden (LB) were found, representing 20·3% of the female population. One had low prevalence across all diseases and risk factors (LB1) and two others were characterised by high prevalence of smoking and either alcohol or drug misuse or asthma alone (LB2 and LB3). Of three cardiometabolic (CM) clusters (18·8% of females), one had very high prevalence across all cardiovascular, kidney and metabolic conditions along with respiratory disease and gout (CM1), another was characterised by a high prevalence of cardiometabolic and neurological conditions including dementia and Parkinson Disease (CM2), and a third had high prevalence of ischemic heart disease and renal disease but lower prevalence of heart failure (CM3). All cardiometabolic clusters had lower prevalence of mental health conditions. Overall, 35·6% of females were in five subtypes of mental health clusters, one with marked high prevalence (MH1) of mental health conditions, smoking and drug or alcohol misuse, one with high prevalence of multiple sclerosis and other neurological conditions (MH2), one with higher prevalence of eating disorders and lower prevalence of cardiovascular disease and risk factors (MH3), one with relatively intermediate prevalence of anxiety depression and a range of diverse conditions (MH4), and finally another by higher prevalence of inflammatory bowel disease (IBD) (MH5). Finally, three mixed condition clusters (21·3% of females) were characterised by cancer, dementia, osteoarthritis and osteoporosis (MIX1), dementia, hearing impairment, osteoporosis and multiple sclerosis (MIX2), and cancer, osteoarthritis and Meniere’s Disease (MIX3).

In males (Figure 3), 14·6% were assigned to one of three low disease burden clusters, one being low in all conditions except congenital disease and melanoma (LB1), another characterised by low prevalence of all conditions (LB2), and another with a high prevalence of asthma alone (LB3). Overall, 34·6% of males were assigned to one of six cardiometabolic clusters. One had higher prevalence of diabetes, gout and obesity (CM1) and another had higher prevalence of diabetes, hypertension and Chronic Kidney Disease (CKD) (CM6). The remaining four cardiometabolic clusters (CM2-CM5) had high prevalence of cardiovascular diseases. Of these, CM2 had particularly high prevalence of cardiovascular conditions, as well as Chronic Obstructive Pulmonary Disease (COPD). CM4 was also characterised by a high prevalence of cardiovascular diseases, but in contrast to CM2, a low prevalence of heart failure and COPD. CM3 and CM5 had an intermediate prevalence of cardiovascular conditions, cancer, dementia and Parkinson’s disease, but CM3 had high prevalence of CKD, COPD, and other neurological diseases which were not higher than expected in CM5. As in females, the cardiometabolic clusters had lower prevalence of mental health conditions. Three mental health clusters (MH1, MH2 and MH3, representing 19·9% of males) each had a high prevalence of anxiety, depression, drug or alcohol misuse and smoking, but MH2 also had high prevalence of neurological conditions, and MH3 had high prevalence of schizophrenia, bipolar disorder and chronic liver disease. Three mixed condition clusters (21·5% of males) were found: one including cancer, respiratory disease and infectious diseases (MIX1), another including gout, osteoarthritis and smoking (MIX2), and another including connective tissue disease, gout and osteoarthritis (MIX3). Finally, 9·5% of males were assigned two respiratory clusters: one characterised by respiratory diseases including asthma, bronchiectasis and COPD (RES1) and another which also included connective tissue disease and IBD (RES2).

### Demographic differences between clusters

Between genders, clusters within the broad groupings correlated well, for example, cardiometabolic or respiratory clusters (Supplementary Figure 4). In some cases, a single cluster in one gender was similar to multiple clusters in the other. For example, the female low burden clusters were similar to male low burden clusters but also shared similarities with male mental health and mixed disease clusters.

The age distributions within each cluster were relatively wide, but with notable, differences in the age distributions between clusters. People assigned to cardiometabolic clusters were older (according to their age at their last record) than those in the low disease burden or mental health clusters (Table 2 and Supplementary Figure 3). The mixed and respiratory clusters had more uniform age distributions reflecting a wider age range of individuals within these clusters compared to other clusters (Table 2 and Supplementary Figure 3).

**Table 2:** Description of patient clusters, size (N) and median age (age of individuals at the time of their last record), and follow-up time in female and male CPRD participants.

| Cluster | Cluster description | N (%) | Median age (IQR), years | Median follow-up (IQR), years |
| --- | --- | --- | --- | --- |
| **Females** |  |  |  |  |
| **Low disease burden (LB)** |  | 650,897 (20.3) | 57 (49 – 70) | 27 (19 – 38) |
| LB1 | Low disease burden and low prevalence of risk factors | 291,665 (9.1) | 57 (48 – 70) | 28 (19 – 39) |
| LB2 | Low disease burden, high prevalence of smoking, and asthma | 190,005 (5.9) | 59 (50 – 70) | 27 (18 – 39) |
| LB3 | Low disease burden, high prevalence of smoking/ alcohol or drug misuse | 169,227 (5.3) | 57 (49 – 68) | 27 (18 – 37) |
| **Cardiometabolic (CM)** |  | 601,704 (18.8) | 83 (74 – 89) | 30 (21 – 42) |
| CM1 | Multiple cardiovascular conditions, COPD, CKD, hypertension, diabetes, gout, dementia | 240,443 (7.5) | 85 (78 – 90) | 29 (18 – 45) |
| CM2 | Multiple cardiovascular conditions, dementia, Parkinson's disease | 194,004 (6.1) | 85 (76 – 91) | 28 (16 – 43) |
| CM3 | Ischemic heart disease, hypertension, CKD, arrhythmia | 167,257 (5.2) | 76 (66 – 85) | 29 (18 – 44) |
| **Mental health (MH)** |  | 1,138,738 (35.6) | 60 (51 – 74) | 30 (21 – 42) |
| MH1 | Multiple mental health conditions, high prevalence of smoking/ alcohol or drug misuse | 246,272 (7.7) | 64 (54 – 76) | 30 (21 – 42) |
| MH2 | Multiple mental health conditions, epilepsy, multiple sclerosis and other neurological conditions, smoking & obesity | 240,490 (7.5) | 67 (56 – 79) | 34 (24 – 46) |
| MH3 | Eating disorders, several mental health conditions, low prevalence of cardiovascular conditions, and risk factors | 239,858 (7.5) | 50 (45 – 56) | 26 (18 – 36) |
| MH4 | Anxiety, depression and mixed conditions of intermediate prevalence | 226,129 (7.1) | 66 (55 – 78) | 31 (22 – 44) |
| MH5 | Anxiety, depression, and IBD | 185,989 (5.8) | 60 (51 – 74) | 31 (23 – 42) |
| **Mixed (MIX)** |  | 683,135 (21.3) | 72 (60 – 83) | 29 (19 – 43) |
| MIX1 | Cancer, dementia, osteoarthritis, osteoporosis | 250,093 (7.8) | 76 (67 – 85) | 28 (17 – 43) |
| MIX2 | Dementia, hearing impairment, osteoporosis, multiple sclerosis | 241,945 (7.6) | 69 (57 – 81) | 31 (21 – 44) |
| MIX3 | Cancer, osteoarthritis, Meniere's disease | 191,097<br>(6·0) | 68<br>(56 – 80) | 28<br>(18 – 41) |
| **Respiratory (RES)** |  | 126,756<br>(4·0) | 64<br>(53 – 76) | 29<br>(20 – 42) |
| RES1 | Asthma, COPD, IBD, and connective tissue disease | 126,756<br>(4·0) | 64<br>(53 – 76) | 29<br>(20 – 42) |
| **Males** |  |  |  |  |
| **Low disease burden** |  | 386,484<br>(14·6) | 57<br>(49 – 68) | 28<br>(18 – 41) |
| LB1 | Low disease burden and low prevalence of risk factors, except congenital disease, and melanoma | 140,398<br>(5·3) | 57<br>(50 – 67) | 30<br>(20 – 43) |
| LB2 | Low disease burden | 129,687<br>(4·9) | 59<br>(50 – 70) | 27<br>(18 – 39) |
| LB3 | Low disease burden, high prevalence of asthma | 116,399<br>(4·4) | 56<br>(48 – 67) | 27<br>(17 – 40) |
| **Cardiometabolic** |  | 914,067<br>(34·6) | 74<br>(64 – 83) | 27<br>(17 – 41) |
| CM1 | Diabetes, gout, obesity | 209,963<br>(7·9) | 65<br>(55 – 76) | 29<br>(20 – 42) |
| CM2 | Multiple cardiovascular conditions, COPD, CKD, diabetes, gout | 170,306<br>(6·4) | 80<br>(72 – 86) | 28<br>(18 – 42) |
| CM3 | Multiple cardiovascular conditions, cancer, COPD, CKD, dementia, Parkinson's disease | 155,216<br>(5·9) | 81<br>(72 – 87) | 28<br>(17 – 42) |
| CM4 | Multiple cardiovascular conditions (except heart failure), CKD, diabetes | 154,298<br>(5·8) | 71<br>(63 – 79) | 27<br>(18 – 41) |
| CM5 | Multiple cardiovascular conditions, cancer, diabetes, dementia, Parkinson's disease | 113,398<br>(4·3) | 77<br>(68 – 85) | 21<br>(12 – 33) |
| CM6 | Diabetes, hypertension, CKD | 110,886<br>(4·2) | 69<br>(60 – 77) | 26<br>(17 – 40) |
| **Mental health** |  | 525,461<br>(19·9) | 59<br>(51 – 70) | 29<br>(20 – 41) |
| MH1 | Multiple mental health conditions, high prevalence of smoking/ alcohol or drug misuse | 204,626<br>(7·7) | 58<br>(50 – 68) | 28<br>(19 – 40) |
| MH2 | Multiple mental health conditions, drug or alcohol misuse, epilepsy, and other neurological conditions, smoking | 189,899<br>(7·2) | 66<br>(56 – 76) | 31<br>(22 – 44) |
| MH3 | Multiple mental health conditions, including schizophrenia, high prevalence of smoking/ alcohol or drug misuse, and chronic liver disease | 130,936<br>(4·9) | 54<br>(47 – 62) | 27<br>(18 – 39) |
| **Mixed** |  | 567,472<br>(21·4) | 65<br>(54 – 76) | 28<br>(19 – 41) |
| MIX1 | Cancer, infectious disease, and respiratory disease | 241,920<br>(9·1) | 71<br>(59 – 81) | 30<br>(20 – 44) |
| MIX2 | Gout, osteoarthritis, smoking | 166,641<br>(6·3) | 60<br>(51 – 72) | 28<br>(19 – 40) |
| MIX3 | Connective tissue disease, gout, osteoarthritis | 158,911<br>(6·0) | 62<br>(52 – 72) | 26<br>(17 – 38) |
| **Respiratory** |  | 251,766<br>(9·5) | 63<br>(52 – 74) | 29<br>(20 – 42) |
| RES1 | Asthma, bronchiectasis, and COPD | 132,836<br>(5·0) | 63<br>(53 – 74) | 29<br>(19 – 42) |
| RES2 | Asthma, bronchiectasis, COPD, IBD, and connective tissue disease | 118,930<br>(4·5) | 62<br>(52 – 74) | 29<br>(20 – 42) |

Abbreviations: CKD = Chronic Kidney Disease; COPD = Chronic Obstructive Pulmonary Disease; IBD = Inflammatory Bowel Disease

There were also significant differences in ethnicity and deprivation distributions across clusters (Figures 4 and 5, Supplementary Table 5). For example, within the male cardiometabolic clusters, distinct subgroups emerged: CM2 had an overrepresentation of white individuals not living in deprived areas, CM1 was predominantly composed of South Asian individuals not living in deprived areas and CM6 had a higher proportion of Black individuals living in deprived areas. Similarly, notable disparities were observed in respiratory clusters (RES1 in females and RES2 in males), which were overrepresented by non-white individuals living in areas with low deprivation. We also found that the low disease burden clusters in both males and females were underrepresented by people living in the most deprived areas.

**Figure 4:**
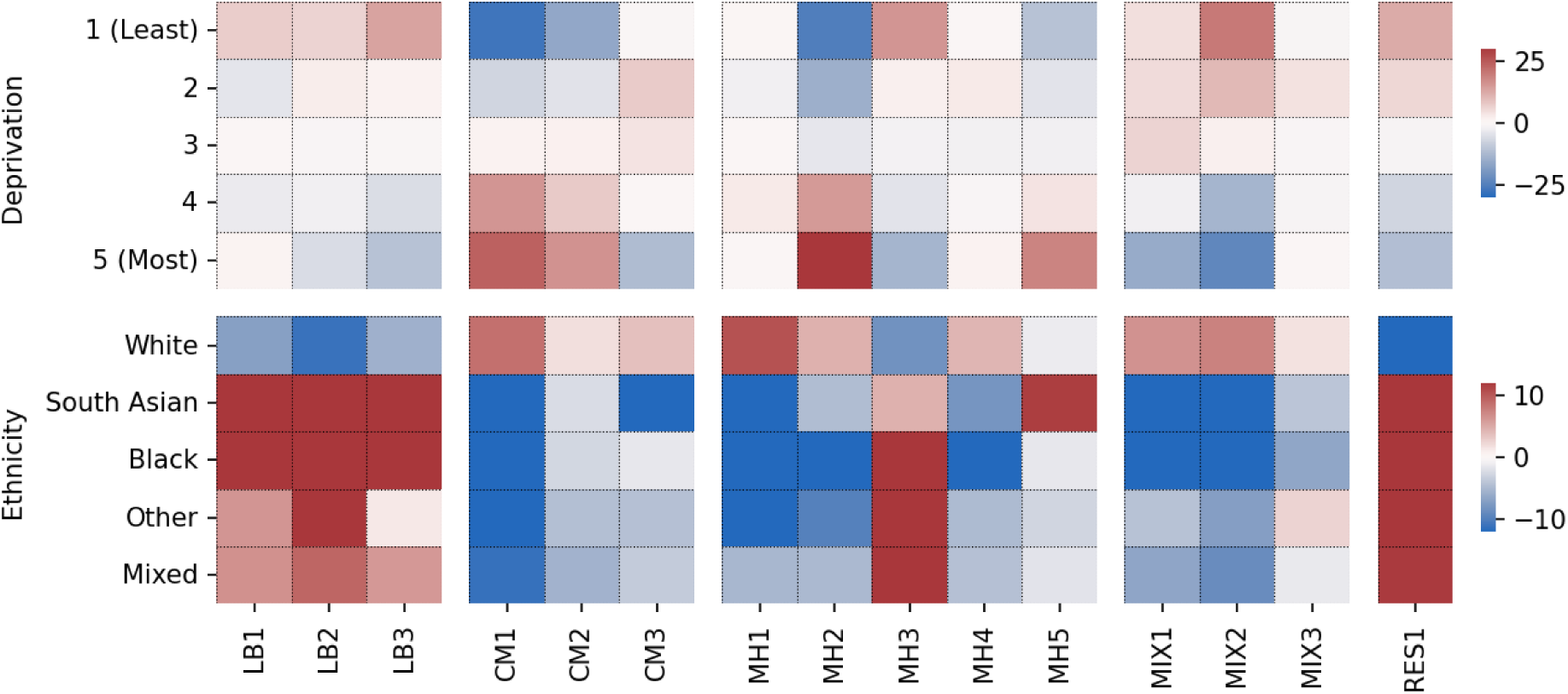
Z-scores differences in observed vs expected frequency in ethnicity and deprivation distributions across clusters in females. LB: low burden, CM: cardiometabolic, MH: mental health, MIX: mixed, RES: respiratory

**Figure 5:**
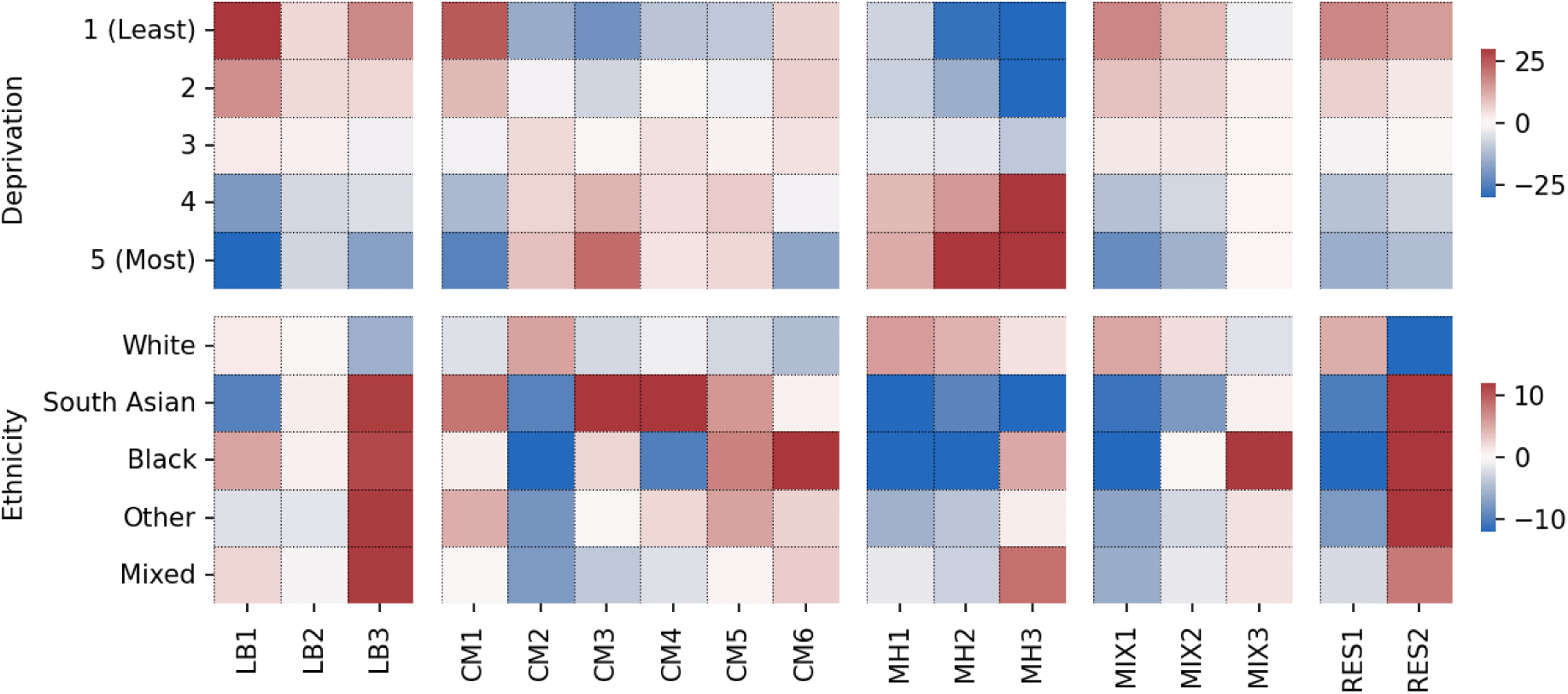
Z-scores differences in observed vs expected frequency in ethnicity and deprivation distributions across clusters in males. LB: low burden, CM: cardiometabolic, MH: mental health, MIX: mixed, RES: respiratory

## Discussion

We analysed a nationally representative population of 5,846,480 million people in the UK, grouping their individual trajectories based on the chronological order of disease development. A key innovation of our method is the use of an LLM to cluster patients rather than diseases, leveraging the full spectrum of clinical information – including prescriptions, diagnoses, and laboratory tests – representing a major advance on earlier work that has primarily mapped disease co-occurrence cross-sectionally. By employing an advanced transformer architecture (EHR-DeBERTa), our approach facilitates a dynamic and more personalised understanding of MLTC patterns, resulting in interpretable gender-specific patient clusters. This approach has significant applications in identifying shared disease pathways, enhancing risk stratification, and optimizing healthcare delivery and management strategies.

We identified five main groups of clusters in both female and male patients: clusters of relatively young and healthy individuals, clusters of middle-aged individuals dominated by mental health conditions, clusters of older individuals with cardiometabolic comorbidities, clusters of patients with respiratory diseases and comorbidities and clusters of patients with mixed conditions spanning different body systems across a wider age range. Within each broad group, there were different cluster subtypes representing distinct patient trajectories including recognisable co-occurrence of linked conditions as well as other less well studied disease combinations. These findings highlight how MLTC manifests across age groups, for example, with mental health conditions being particularly prevalent in middle age and cardiometabolic diseases become more dominant in older populations. This age-related variation highlights the need for tailored healthcare strategies that address the evolving burden of MLTCs over the life course and emphasizes the importance of early interventions before disease has occurred, for example, to prevent transitions from a low disease burden cluster to a mental health cluster in midlife or a cardiometabolic cluster later in life.

In females, mental health clusters were more frequent, accounting for 28% of females versus only 13% of males, reflecting the higher prevalence and risk of mental health – particularly depression and anxiety – in females compared to males.^29^ In both genders, we identified a highly comorbid cluster with multiple mental health conditions, co-occurring with diabetes, obesity, and neurological conditions including Parkinson’s Disease and multiple sclerosis.^30,31^ These findings reinforce existing epidemiological evidence on bidirectional associations between these conditions^32^ and further stresses the need for integrated management approaches that address both mental and physical health within a unified care framework.

We observed significant variation in the prevalence of cardiometabolic conditions across clusters, suggesting that these conditions play a dominant role in distinguishing patient phenotypes. Cardiometabolic clusters were more prevalent in males compared to females and females in cardiometabolic clusters tended to be older than their male counterparts, consistent with literature.^33,34^ However, the composition of clusters showed notable similarities between genders. In both males and females, we observed a highly comorbid cluster of cardiovascular, metabolic and renal comorbidities, often co-occurring with other linked conditions such as gout, COPD, and neurodegeneration. This underscores the well-recognised bidirectional association between the dysfunction of the heart, kidneys and metabolic system, and their systemic multi-organ consequences.^35^ The co-occurrence of neurodegeneration highlights an important heart-brain-liver axis which is less well-characterised. As more therapeutic options emerge, it is crucial to develop integrated treatment strategies that maximize multi-organ benefits and establish shared guidelines for managing these interconnected conditions. Simultaneously, a multi-organ approach to disease prevention could enable to early intervention as abnormalities in any of these organs may serve as early indicators of disease progression, offering opportunities for timely intervention, and improved patient outcomes. Interestingly, mental health and cardiovascular clusters were mutually exclusive: clusters with a high prevalence of cardiometabolic conditions had a lower prevalence of mental health conditions and vice versa. This contrasts with strong epidemiological evidence of increased incidence of cardiovascular disease in those with pre-existing mental health conditions, particularly psychotic disorders.^36^ Possible explanations include the younger age of those in the mental health clusters, who may not yet have developed cardiovascular disease, and the older age of those in the cardiometabolic clusters, mental health conditions may be underdiagnosed due to stigma and barriers to accessing support.^37^

Other multi-organ effects were evident from the patient clustering. For example, the clustering of patients with IBD with COPD and other respiratory conditions. These conditions may share similar underlying pathways such as infection, immune activation, inflammation, and shared microbiota effects which may lead to common treatments and monitoring of patients with IBD for the potential of developing respiratory disease.^38^ Similarly, the association between osteoporosis and dementia raises the possibility of an underlying biologic mechanism linking bone loss and cognitive decline, especially in females.^39^

The identified clusters of patients differed substantially based on deprivation and ethnic diversity. Within each broader patient group, individuals with similar ethnic backgrounds and socioeconomic status tended to cluster together. For example, we identified distinct male cardiometabolic clusters with an overrepresentation of White individuals (CM2), South Asian individuals (CM1), and Black individuals (CM6), with the latter two clusters having a lower median age than CM2. We also found that low disease burden clusters were underrepresented by people living in more deprived areas. Several conclusions can be drawn. Firstly, MLTCs are a widespread issue across the population and although not confined to populations with specific ethnic or socioeconomic backgrounds, people in low disease burden clusters are more likely to be from less deprived areas. Secondly, individuals with similar socioeconomic status and ethnic backgrounds may experience comparable trajectories of comorbidities, reflecting shared risk factors within these groups. Thirdly, clusters including people living in areas of greater deprivation and from non-White ethnic backgrounds tend to be younger, possibly indicating earlier onset of MLTCs in these subgroups. Previous research has also highlighted that people living in more deprived areas have a faster accumulation of conditions and higher mortality rates.^40^ Collectively, our findings emphasise that tailored healthcare strategies are essential to address the diverse needs of different population groups in addition to integrated approaches to managing MLTC.

Traditional clustering approaches in healthcare often focus on either patient characteristics or clinical outcomes.^19^ LLMs offer an opportunity to combine both, by fine-tuning on outcomes such as mortality or hospital admission. This could enable the generation of clusters that not only reflect patient characteristics but also predict downstream risks. Future research could investigate the relationship between model self-attention and cluster assignment to identify sequences of diseases which are at higher risk of adverse health outcomes. Simulated interventions can also be explored by modifying patient histories (e.g., adding medication at a specific time point or delaying a diagnosis), informing personalised lifestyle, or treatment recommendations and drug repurposing. Fine-tuned models could also support clinical decision-making by providing the probability of a missed diagnosis or the potential effect of an interventions for each patient. Another strength of this study is the use of a large, contemporary dataset which is representative of the English population.^20^ An advantage of primary care data is that historic diagnoses can be entered retrospectively with the date of diagnosis, capturing a more complete view of a person’s disease history than is often available in hospital records.

Some limitations also need to be acknowledged. Although the prevalence of many common diseases in CPRD is similar to other national data sources,^21^ for conditions such as cancer, diagnosis recording is less complete than estimates derived from gold standard sources such as cancer registry data.^22^ The presence or absence of a diagnostic code in GP EHR data is also influenced by factors independent of a person’s health, such as GP practice organisational coding policies and financial incentives, potentially biasing model learning towards conditions which are recorded more frequently.^41^ We summarised continuous diagnostic tests as binary outcomes (e.g., abnormal C-reactive protein levels) which may result in loss of information. Future work could explore use of additional embedding layers that can provide information on continuous variables, such as blood pressure readings. We utilised a hard clustering method, using a robust method to ensure the optimal cluster number and evaluated the quality of the clustering with multiple methods. However, the optimal number of clusters is difficult to define as there may be multiple optima with similar performance. Future studies employing soft-clustering methods may allow for more flexible identification of patient clusters. We presented the single diseases that were prevalent within each cluster. While we presented individual diseases prevalent in each cluster, the underlying embeddings also captured disease sequences over time. Future work could explore whether common diseases trajectories differ between clusters that otherwise appear similar.

We demonstrated the potential of an LLM-based clustering pipeline to identify gender-specific clusters of patients and to provide a more nuanced and personalised understanding of MLTC patterns by leveraging longitudinal EHR data. Our findings also indicate valuable opportunities to explore shared disease mechanisms and potential drug repurposing applications. Cardiometabolic and mental health conditions emerged as the dominant drivers of distinct patient phenotypes, with notable differences in males and females. We observed clear differences in MLTC composition across the lifespan, with mental health conditions more dominant in middle age and cardiovascular diseases more prevalent in older adults, and with unique MLTC clusters shaped by differences in ethnic and socioeconomic backgrounds. Collectively, these findings underscore the need for integrated health services and interventions, which are patient-centred, rather than disease-centred. Future work should focus on refining clustering methodologies, incorporating clinical outcomes, and exploring the utility of clusters to inform real-world models of care in healthcare settings. Ultimately, integrating LLM-driven patient clustering into clinical practice could enhance risk stratification, enable early detection of conditions and improve long-term health outcomes for people with MLTC.

## Data Availability

All data are available upon request and accepted protocol by CPRD.

## Acknowledgements

The study and AS were funded by the NIHR Imperial College London Biomedical Research Centre.

## Contributors

AS and IT conceived the project. AS designed the study and did the analysis. AS and TB interpreted the data and wrote the original draft. IT supervised the work. All authors reviewed and interpreted the results, commented on the paper, contributed to revisions, and read and approved the final version. IT had final responsibility for the decision to submit for publication. Only individuals involved in directly managing or analysing the data are allowed to access the data, hence only AS and TB had access to the raw data. All authors had access to aggregated data and statistical outputs. Data management was provided by the Big Data and Analytical Unit (BDAU) at the Institute of Global Health Innovation (IGHI), Imperial College London.

## Appendix

## Supplementary Methods

### Constructing patient sequences

Clinical diagnoses included all conditions encoded at the highest level of the ICD-10 hierarchy across all diseases or health conditions chapters (Chapters A to S), and all SNOMED-CT codes which mapped to an ICD-10 code within these chapters. Primary care records of interest were mapped to ICD-10 codes prior to inclusion chronologically in each patient sequence and records with no mapping to an ICD-10 code were excluded. ICD-10 codes that occurred in less than 100 individuals within the entire study were excluded to reduce vocabulary size. Symptoms and laboratory tests were included chronologically using ICD-10 Section R and a manually curated list of SNOMED-CT codes which encode indicators of abnormal test results. Products within the prescription records were first mapped to medication groups based on the primary function of the product (Supplementary Table 3) and subsequently included chronologically in each patient sequence resulting in a final vocabulary size of 3776 tokens. To reduce the chance of continuous prescriptions dominating each patient sequence, only the first instance of mapped medications prescribed monthly were included in the chronological sequences with a six-month gap in prescription required before the re-addition of the medication code to the sequence. For mapped medications not prescribed monthly, all prescription events were included chronologically in each patient sequence. Each event was also paired with additional information related to the event: the age of the individual at the point of the event, the calendar year that the event occurred and the visit number for the patient to a medical professional. The visit number was defined incrementally starting at the first medical event for each patient, with events occurring within the same month considered the same visit.

### Model training

Our ELECTRA style pre-training involved training two models simultaneously using a replaced token detection (RTD) task by firstly masking 15% of clinical events within a sequence and utilising an EHR-DeBERTa model, named the generator, to predict the masked code. The sequence created by the generator with the masked events replaced was then fed into a second EHR-DeBERTa model, called the discriminator, which predicted if each code in the sequence was part of the original sequence or if it has been replaced by the generator. Both models are updated at the same time, utilising a dis-entangled clinical event embedding between the discriminator and generator implemented in the DeBERTa v3 procedure to improve model training. We used the same hyperparameters as the small DeBERTa v3 model consisting of 6 layers in the discriminator and 3 layers in the generator, as described by the original authors.^23^ In doing this pre-training procedure, the generator builds up a vector representation of each clinical event within its vocabulary based on each clinical events relationship with other events and their interactions with age, sex, calendar year and visit number. The models were trained for a total of 100 epochs to ensure convergence using dynamic masking as suggested by RoBERTa with a 15% of masking rate.^42^ Training took a total of one week using a single NVIDIA Quatro RTX 8000.

We subsequently fine-tuned the pre-trained EHR-DeBERTa generator using DiffCSE which is an equivariant contrastive learning method that ensures the generated patient representations are insensitive to certain types of event alterations within patient sequences, such as swapping a single clinical event to an event which has almost no difference to patient trajectory, and sensitive to other types of event alterations that would cause a drastically different trajectory, considered “harmful” alterations. This was performed using the pretrained EHR-DeBERTa generator model with temperature = 0.05 and lambda = 0.005, as suggested by the original authors, for 5 epochs.

### Cluster specific disease frequency – inverse patient frequency (c-DF-IPF)

We generated cluster specific weighted frequencies by adapting the algorithm for cluster specific term frequency – inverse patient frequencies. In this context, a patient’s medical history is treated as a document and the population is treated as the corpus and converted into a disease presence or absence (binary variable) matrix.

We initially calculate the global importance of diseases across the population using the entire corpus. Let *P* represent the entire patient cohort, we apply DF-IPF:

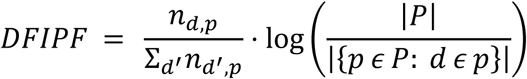

where *n*_*d*,*p*_ is the presence of disease *d* in patient record *p* and 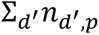 is the total number of diseases within the patient’s record.

To calculate cluster specific importance, we averaged the DF-IPF of individuals within each cluster and scaled these using cluster specific inverse patient frequencies, *IPF*_*i*_:

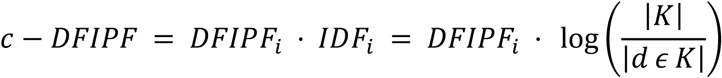

where *DFIPF*_*i*_ represents the average cluster importance of cluster *i*, |*K*| is the total number of clusters and |*d ∈ K*| is the number of clusters disease *d* is present within.

